# A Comparison of Representation Learning Methods for Medical Concepts in MIMIC-IV

**DOI:** 10.1101/2022.08.21.22278835

**Authors:** Xuan Wu, Yizheng Zhao, Yang Yang, Zhangdaihong Liu, David A. Clifton

## Abstract

**Objective:** To compare and release the diagnosis (ICD-10-CM), procedure (ICD-10-PCS), and medication (NDC) concept (code) embeddings trained by Latent Dirichlet Allocation (LDA), Word2Vec, GloVe, and BERT, for more efficient electronic health record (EHR) data analysis.

**Materials and Methods:** The embeddings were pre-trained by the four aforementioned models separately using the diagnosis, procedure, and medication information in MIMIC-IV. We interpreted the embeddings by visualizing them in 2D space and used the silhouette coefficient to assess the clustering ability of these embeddings. Furthermore, we evaluated the embeddings in three downstream tasks without fine-tuning: next visit diagnoses prediction, ICU patients mortality prediction, and medication recommendation.

**Results:** We found that embeddings pre-trained by GloVe have the best performance in the downstream tasks and the best interpretability for all diagnosis, procedure, and medication codes. In the next-visit diagnosis prediction, the accuracy of using GloVe embeddings was 12.2% higher than the baseline, which is the random generator. In the other two prediction tasks, GloVe improved the accuracy by 2%-3% over the baseline. LDA, Word2Vec, and BERT marginally improved the results over the baseline in most cases.

**Discussion and Conclusion:** GloVe shows superiority in mining diagnoses, procedures, and medications information of MIMIC-IV compared with LDA, Word2Vec, and BERT. Besides, we found that the granularity of training samples can affect the performance of models according to the downstream task and pre-train data.

## 1 Introduction

In the EHR systems, medical codes in diagnosis, procedure and medication contain rich medical information and abundant hidden knowledge about disease/therapeutic characteristics. Though the codes in different EHR system has distinct distribution due to different demographic of patient population, the hidden knowledge between these medical codes are universal. Existing EHR mining research [7, 31, 41] widely use predefined (also known as embedding) medical codes to capturing the hidden correlation between codes and enhancing various clinical tasks, such as diagnosis prediction and mortality prediction.

With the widespread deployment of EHR systems, clinical datasets such as MIMIC-IV [19] may store rich patient information while retaining their multimodality and complex structures. Well-structured code systems in EHR often record patient information such as diagnoses, procedures, and medications. A medical code is equivalent to a ‘word’ in natural language; all codes that occurred during a patient’s hospital visit naturally constitute one ‘sentence’; and a ‘document’ is equivalent to all hospital visits of a patient sorted by visiting time. The major inadequacy of this analogy is that each visit is a time-irrelevant set of codes. In other words, unlike natural language, the ‘words’ in each ‘sentence’ here do not appear in any particular order. As a result, order-insensitive sequence models based on the distributional hypothesis (LDA [2], Word2Vec [25], and GloVe [33]) are suitable for modelling medical codes. A more complex language model, BERT is also analyzed in this paper for comparison.

### 1.1 Word Embedding

In natural language processing (NLP), the semantic and syntactic features of words in unlabeled text data are expected to be fully captured in a low dimensional space through distributional semantics word embedding. Under distributional hypothesis [15], two semantically similar words are assumed to occur in a similar context, which means having similar word co-occurrence. Word2Vec [25] defines the context of the target word as the co-occurrence between target word and the words in the fixed-size window around it. As Word2Vec only considers the local cooccurrence, GloVe [33] utilizes global co-occurrence to capture explicitly defined sub-linear relationships. However, the drawback of these early word embeddings is that they represent all senses of a word into one embedding, while words are in multiple senses, in reality, e.g. “good” and “cheerful” are synonyms, but mean different in sentences “This restaurant has a good reputation.” and “A great many people would agree.”. Then many sense embedding works are proposed to give representations in different senses to a word in supervised or unsupervised ways. Some unsupervised sense embeddings works [28, 23] are based on LDA [2], a probabilistic topic model that can model the topic distribution over documents. In recent years, contextual embedding methods arises to give a solution that covers many aspects (e.g. sense representation, synonymy/antonymy, and hypernymy/hyponymy) [37]. The successful contextual embedding method BERT [10] learns contextual word embeddings based on bidirectional transformer architecture with the masked language model (MLM).

### 1.2 Medical Codes Representation Learning

Since the arising of deep learning, representation learning of medical codes becomes crucial for clinical prediction. Early works learn visit/patient representations [6, 6, 8, 29, 27, 4] utilizing deep learning methods such as recurrent neural networks, convolutional neural networks or stacked denoising autoencoders on longitudinal EHR data. These methods demonstrates the effectiveness of learning distributional representation for clinical prediction, but the representations are prediction-task-guided. For adaptation of different patient distribution in different EHR dataset, many works turn to utilizing language model to embed medical concepts. GRAM [7] learns ICD-9 disease code embeddings using GloVe and shows a considerable improvement in sequential diagnosis predictions; G-BERT [38] pre-trains on ICD-9 disease codes for diagnoses and ATC codes for medications together with a modified MLM objective where the loss combines these two modalities of codes; BEHRT [22] embeds caliber codes for diagnoses with only MLM pre-training objective and achieved better results than visit and patient representation works [8, 27] in next visit prediction; Med-BERT [34] embeds ICD-9 and ICD-10 codes for diagnoses to predict the prolonged length of stay in hospital task with MLM pre-training. Recently, some works investigated the performance of other unsupervised NLP models in embedding medical concepts. Finch et al. [13] shows the clustering capability of ICD-10 disease codes with Word2Vec embeddings. Nuria et al. compared LDA and PLDA (supervised version of LDA) [21] ICD-10 in Osa dataset and ICD-9 in MIMIC-III dataset. However, for a variety of reasons, most of these works are unable to publish the pre-trained code embeddings. And as far as we know, there is no such comparison between these word embedding methods in longitudinal EHR data. The contribution of this work is twofold: (1) it gives an empirical comparison of four popular code embedding pre-training methods which can be considered as guidance for EHR researchers; (2) we are the first to publish the publicly accessible pre-trained embeddings^1^ for ICD-10 codes using BERT, GloVe, Word2Vec, and LDA based on MIMIC-IV data, which can be used as initialization for any ICD-10 related tasks.

## 2 Method

### 2.1 Comparison Framework

To compare the pre-trained embeddings, LDA, Word2Vec, GloVe, and BERT were assessed in two ways: embedding interpretability and performance in three downstream tasks.

We set our pre-training to be downstream task-independent in order to make the application of the pre-trained embeddings as wide as possible. We note that the pre-training methods are unsupervised/self-supervised learning: the embeddings are learned from the intrinsic information in the data, such as the co-occurrence or the context information, and are not further tuned with the down-stream tasks.

The three modalities of medical concept, ICD-10-CM (Clinical Modification of diagnoses), ICD-10-PCS (Procedure Coding System), and NDC (National Drug Code), may have different compatibilities with LDA, Word2Vec, GloVe and BERT, therefore, were individually trained.Then the medical concept embeddings were fixed (without fine-tuning) and fed into downstream tasks as the input features of the concepts.

We extracted the diagnosis, procedure and medication information from the ‘diagnoses_icd’, ‘procedures_icd’ and ‘prescriptions’ tables under the ‘hosp’ module from the MIMIC-IV (version 1.0) website (https://physionet.org/content/mimiciv/1.0/), respectively. In particular, we focused on the ICD-10 codes for diagnosis and procedure and NDC codes for prescription. The statistical information of the data is depicted in Table. 1. All of the data were processed into two levels of granularities: each patient as a sample and each visit as a sample, as depicted in Figure. 1. When using a patient as a sample, we concatenated all visits of a patient ordered by time to form a ‘sentence’; when a visit serves as a sample, each ‘sentence’ is simply the collection of all codes that occurred during a visit. Notably, the codes within a visit are not ordered by time since this information is not provided in MIMIC. As far as we know, the choice of sample granularity has not been studied under the representation learning of medical concepts.

**Table 1.**
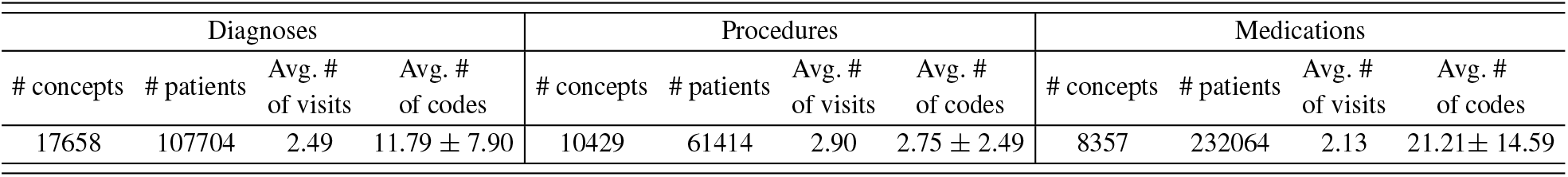
Information on diagnoses, procedures, and medications data. Avg. # of visits is on patients and Avg. # of codes is on visits.

| Diagnoses |  |  |  | Procedures |  |  |  | Medications |  |  |  |
| --- | --- | --- | --- | --- | --- | --- | --- | --- | --- | --- | --- |
| # concepts | # patients | Avg. # of visits | Avg. # of codes | # concepts | # patients | Avg. # of visits | Avg. # of codes | # concepts | # patients | Avg. # of visits | Avg. # of codes |
| 17658 | 107704 | 2.49 | 11.79 ± 7.90 | 10429 | 61414 | 2.90 | 2.75 ± 2.49 | 8357 | 232064 | 2.13 | 21.21 ± 14.59 |

**Figure 1:**
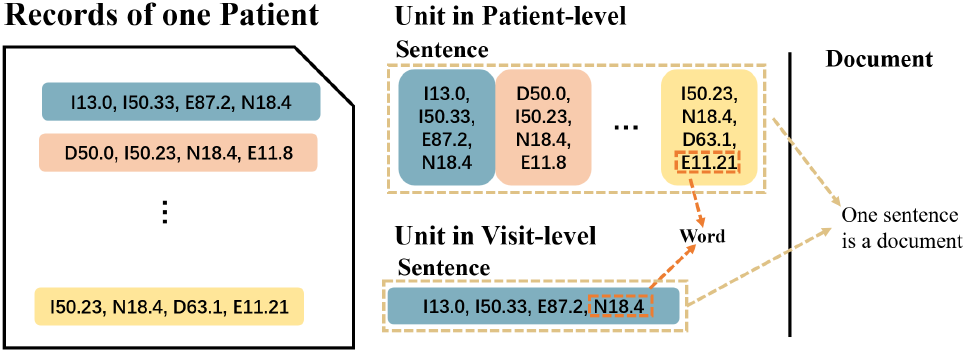
Data format for pre-training in the context of diagnoses data. Blocks with different colors are different visits. The same settings were used for procedures and medication data. A word in diagnoses data is an ICD-10-CM code, while for procedures and medications data is an ICD-10-PCS code, and an NDC code, respectively.

### 2.2 Models

#### 2.2.1 Notation Definition

Let the sets of codes in diagnoses, procedures, and medications data be 𝒞_*D*_, 𝒞_*P*_, and 𝒞_*M*_, respectively. We write 𝒞 as the set of codes when there is no need to specify the modality. 𝒞 is the code vocabulary. A sentence at visit-level is a list constructed by *c* ∈ 𝒞 of the form 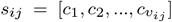, where *v*_*ij*_ is the amount of codes in the *j*th visit of the *i*th patient. While a sentence at patient-level is in the form 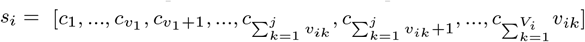, where *V*_*i*_ is the amount of visits of the *i*th patient. The pre-training data consist of a document, which can be defined as *D*. At visit-level, 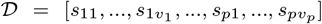, while at patient-level, *𝒟* =[*s*_1_, *s*_2_, …, *s*_*p*_], where *p* indicates the amount of patient.

#### 2.2.2 LDA

Based on De Finetti’s classic representation theorem [9], LDA mixture models capture the exchangeability of both words and documents. LDA assumes that each document is a mixture over latent topics, where each topic is characterized by a distribution over words. A topic distribution *θ*_*i*_ for document *𝒟*_*i*_ is sampled from a Dirichlet allocation *Dir*(*α*). The topic *z*_*i,j*_ for the *j*th word in *𝒟*_*i*_ is sampled from the multinomial distribution *Multinomial*(*θ*_*i*_). Sample 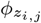 of the *j*th word *c*_*i,j*_ is from the dirichlet distribution *Dir*(*β*). The word *c*_*i,j*_ then is modelled by the multinomial distribution 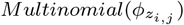. Considering the successful application of LDA in learning phenotypes compared to other topic models, we assume that each dimension of an embedding is a latent topic, and the latent topics can be learned by LDA, so we used the word-topic matrix as the representation of the medical concepts. In this case, the embedding for each word is a vector of the probability in each topic, where the number of topics is the vector dimension. Then, the distribution is learned through variational inference. We implemented LDA using Python library *GenSim* [35], and the inference method is online variational Bayes [17].

#### 2.2.3 Word2Vec

Word2Vec can make use of either continuous bag-of-words (CBOW) [25] or skip-gram [26]. The assumptions are different between CBOW and skip-gram. Skip-gram model assumes that a word can be used to generate its surrounding words, while CBOW assumes that the target word can be generated by its surrounding words. The surrounding words are the words around the target word within a fixed window size. We adopted the skip-gram version of Word2Vec in this work, as our early experiment showed that skip-gram has better performance in this case.

Given the dictionary 𝒞, the training target of skip-gram is to maximize the average log probability:

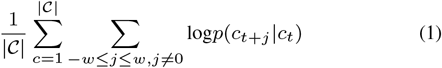

The conditional probability *p*(*c*_*t*+*j*_ |*c*_*t*_) is the probability of the presence of *c*_*t*+*j*_ given the cotext word *c*_*t*_ and is calculated as:

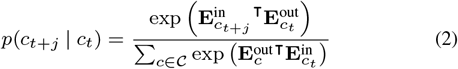

where **E**^in^ and **E**^out^ are the ‘input’ and ‘output’ embeddings of codes. The final embedding for each code is the average between **E**^in^ and **E**^out^.

We used the implementation of Word2Vec provided by Python library *GenSim* with hierarchical softmax and set the window size as 2.

#### 2.2.4 GloVe

While Word2Vec learns the embeddings by mining the local context information, GloVe was proposed to globally compute the word embedding based on the matrix of word-word co-occurrence counts. Instead of considering the local co-occurrence within a fixed-size window, GloVe computes the co-occurrence within a data unit. In our use case, the data unit is the lists of clinical codes at visit/patient level. The distribution of code co-occurrences in GloVe is modelled by a power-law function of the frequency of each code pair. Based on the observation of the co-occurrence probabilities, GloVe models the ratio of co-occurrence probabilities.

#### 2.2.5 BERT

BERT [10] uses a masked language model (MLM) to enable the pretraining of deep bidirectional embeddings, which can give both sentence embeddings and word embeddings. We take diagnosis data as an example and show the structure of BERT in Figure. 2. The pre-training objective of MLM is to predict the randomly replace tokens into ‘[MASK]’ tokens in the sentence with cross-entropy loss. In this way, code embeddings try to learn through their context. We pre-trained the MLM with 15% words masked in a sentence. We only used ‘[MASK]’ tokens but not ‘[CLS]’ and ‘[SEP]’ tokens since only MLM was considered here.

**Figure 2:**
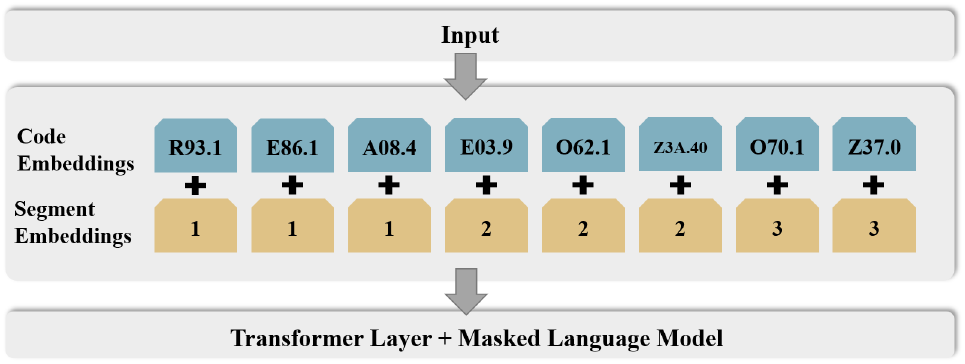
BERT model structure in the context of diagnoses data.

#### 2.2.6 Settings

The model architectures are fixed for the three data modalities, diagnosis, procedure, and medication. To pre-train BERT, we only adopted the patient unit, as the visit unit is too short to show the advantage of BERT. For the other three methods, both of the data granularities were tested. The dimension of the embeddings was fixed as 128 for all models. In the pre-training, the optimizer Adam was employed. The learning rate was tuned as 5e-5, 1e-2, and 2.5e-2 for BERT, GloVe, and Word2Vec respectively. BERT was only pre-trained with MLM, following the setting of Med-BERT [34]. For Word2Vec, we set the window size as 2 with hierarchical softmax. The hyper-parameters in GloVe were set the same as the recommended settings in [33]. For LDA, the dimension of the embeddings was the number of topics.

## 3 Embedding Interpretability

To evaluate the interpretability of the embeddings learned by these models, we used t-SNE [40] to visualize the embeddings in 2D space, and we labelled points according to different standards. For ICD-10-CM codes, we classified points into six typical diseases as specified in Table. 4. While the coding system of ICD-10-CM codes also provides a standard for classifying concepts, so we divided these codes into different categories according to the coding system^2^. For ICD-10-CM codes, we visualize the embeddings with these two grouping standards and show them in Figure. 3. Each ICD-10-PCS code consists of seven characters ^3^. The first character is the ‘section’, and the second through seventh characters mean different things in each section. For ICD-10-PCS, we used the 1st, 2nd, and 3rd characters to classify embeddings, respectively. For medication, we mapped the NDC codes into the Anatomical Therapeutic Chemical (ATC) code system [30], which is a hierarchical classification system that assigns a unique code to each medicine according to the organ system the medicine works on and how it works. We used the top-level ATC as the cluster labels which denote the main anatomical or pharmacological groups.

**Figure 3:**
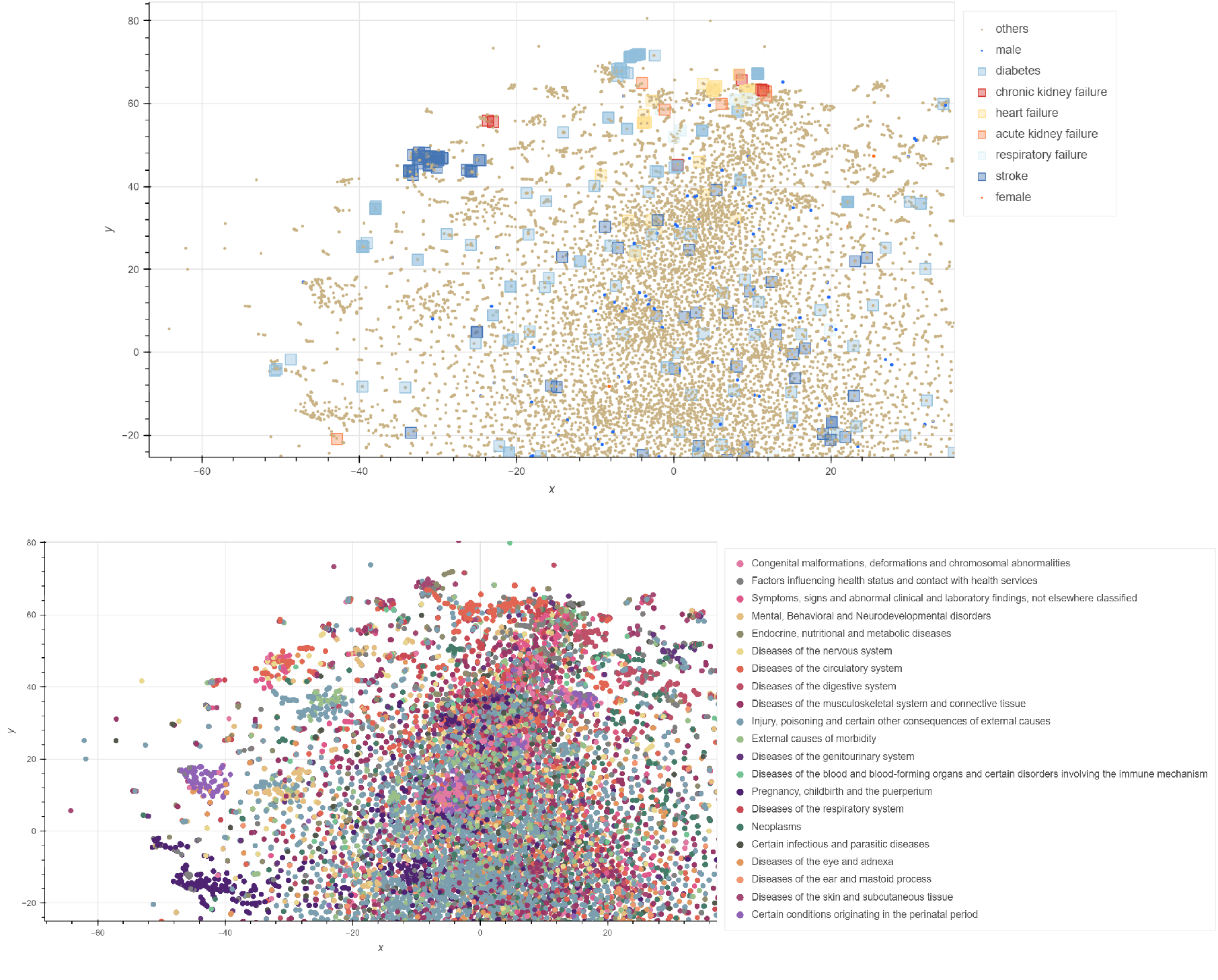
Visualization of GloVe pre-trained ICD-10-CM code embeddings reduced into 2D space by t-SNE, zoomed-in views. The points in these two subfigures are the same, but grouped by different categories.The points upper subfigure are classified into six typical diseases; the lower subfigure is classified according the top level of ICD-10 taxonomy.

More visualizations can be seen in our repository. For diagnoses data, GloVe embeddings did the best in clustering diseases in kidney failure, stroke, and diabetes. LDA and BERT could also cluster chronic kidney failure well. All embeddings trained by the four methods cannot separate between male diseases and female diseases well. For procedure and medication data, embeddings pre-trained by GloVe also gave the best visualization.

To further show the clustering ability of the 4 × 3 sets of embeddings, we used the clustering evaluation metric Silhouette coefficient [36]. The Silhouette coefficient is a value in the range of [-1, 1] measuring how similar points are within a cluster, where a larger value indicates a tighter cluster. Given embeddings and labels, the Silhouette coefficient for an embedding is 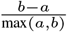, where *a* is the mean distance between the embedding and all other embeddings in the same cluster, and *b* is the distance between the embedding and the nearest cluster that the embedding is not a part of. It is worth noting that all of these four models are not for clustering, so the application of Silhouette coefficient is mainly for the interpretability purpose. We computed the Silhouette coefficient in the original data space (128-d), and used one of the aforementioned labelling systems as clustering labels.

For ICD-10-CM embeddings, the visualization of Silhouette coefficients is shown in Figure. 4 with the top-level categeries of ICD-10-CM. LDA clustered ‘Congenital malformations, deformations, and chromosomal abnormalities’ class well, while GloVe clustered ‘Certain conditions originating in the perinatal period’ better.

**Figure 4:**
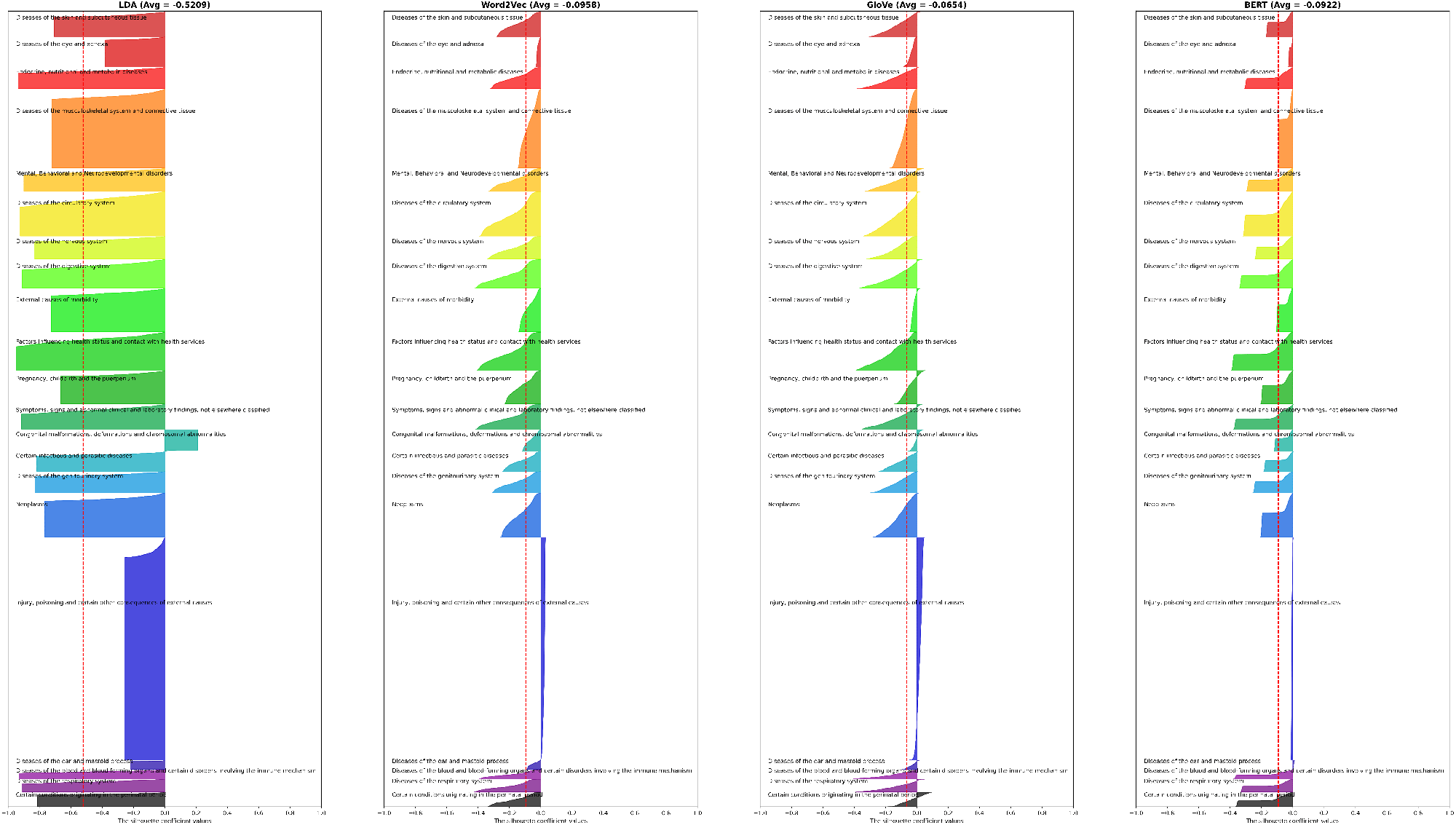
The silhouette coefficient value in clustering ICD-10-CM embeddings. The red dashed line denotes the average silhouette coefficient value.

For ICD-10-PCS embeddings, we used the 2nd character as cluster labels, with results shown in Figure. 5. For most classes, LDA did the worst, for some small clusters, e.g., ‘Breast’, ‘Ear, Nose, Sinus’, ‘Lymphatic System’, LDA could knit them tight.

**Figure 5:**
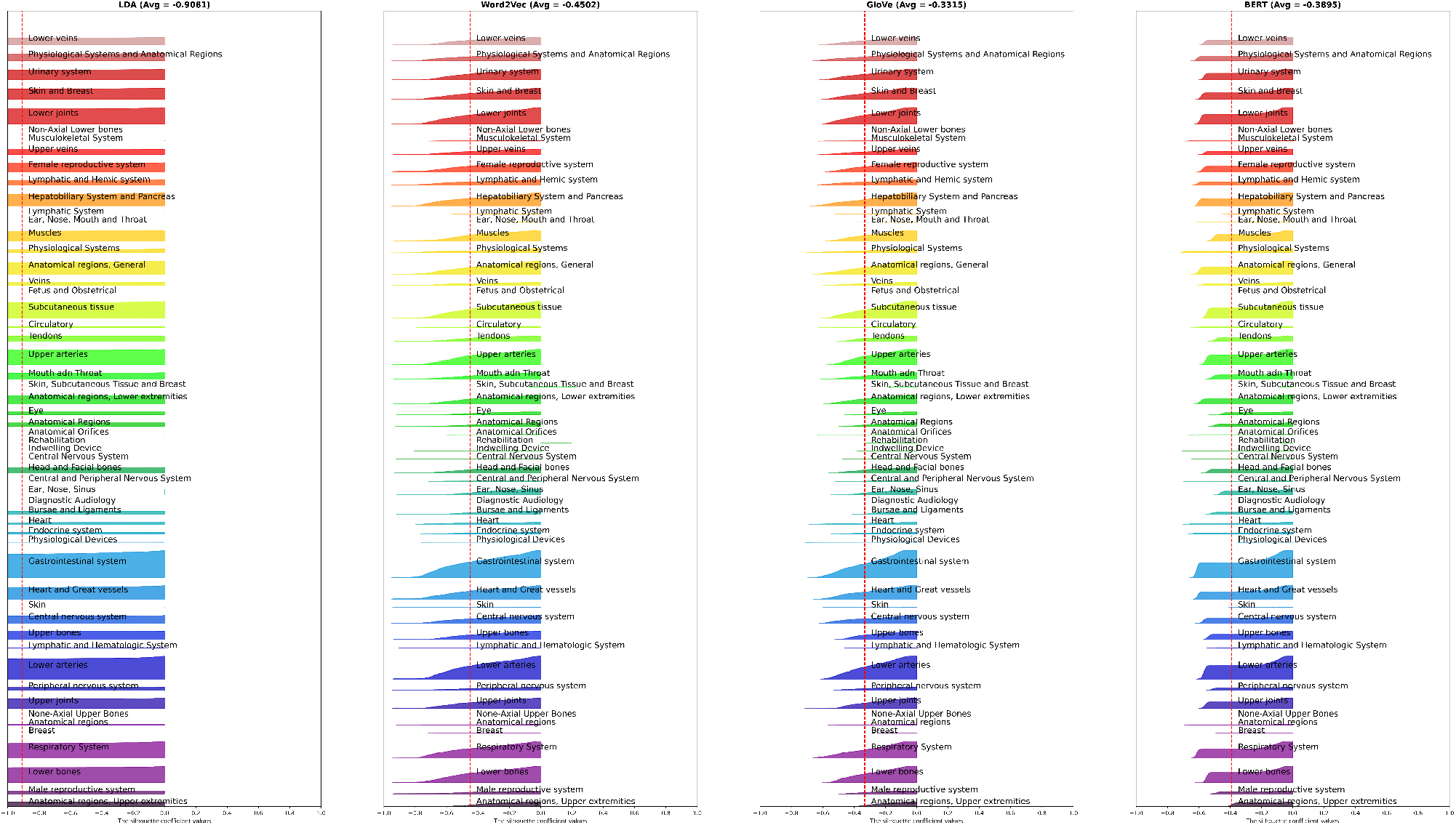
The silhouette coefficient value in clustering ICD-10-PCS embeddings.

For NDC embeddings, in Figure. 6, the ones learned by Word2Vec clustered ‘Antineoplastic and immunomodulating agents’ well. Overall, comparing the average silhouette coefficient, embeddings learned by GloVe displayed better clustering ability than the ones learned by other models.

**Figure 6:**
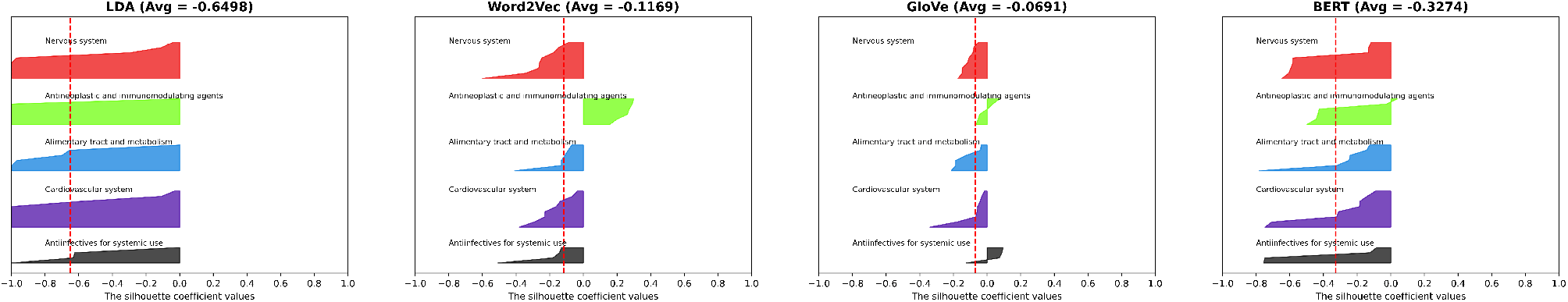
The silhouette coefficient in clustering NDC embeddings.

## 4 EVALUATION

### 4.1 Tasks

To further compare the performance of these embeddings, we evaluated them in three tasks: diagnoses prediction of multiple diseases in the next visit within one month, mortality prediction for intensive care unit (ICU) patients, and medication recommendation. We set the diagnoses prediction as a six-class classification task using diagnoses embeddings, predicting chronic kidney failure, acute kidney failure, heart failure, respiratory failure, diabetes, and stroke for the patients’ next visit. The mortality prediction is a binary classification task predicting whether mortality happens to ICU patients within a month of the current visit using diagnoses/procedures/medication embeddings. The medication recommendation is a regression task that uses the diagnoses/procedures embeddings to predict the medications within the same visit.

Patients with less than 2 visits are omitted in the evaluation. The patients were split into train and test sets with a ratio of 70% and 30%. Then patient-level data were further unwrapped to visit-level since one patient may have multiple visits.

Each training input of these tasks is a set of codes. The pre-trained embeddings for these codes are the feature matrix. To combine these codes’ feature vectors, we use the average of the code embeddings as the feature vector of each input. Except for the four sets of pre-trained embeddings, we also use randomly initialized embeddings to serve as baselines in the three modalities of data. We used four machine learning models — CatBoost [11], Xgboost [5], Random Forest [3] and Multi-layer Perceptron (MLP) [16] for classification (/regression).

#### 4.1.1 Diagnoses Prediction

As we know, multiple organ failure is a severe, life-threatening condition, while stroke and diabetes are correlated to multiple organ failure. The diagnoses prediction in this paper is the multi-classification between multiple organ failure diseases, stroke, and diabetes. This task was conducted on a cohort where patients have more than one visit. We further selected visit pairs with intervals of less than one month, and each visit was labelled with one of the six disease classes mentioned above according to the diagnosis code. Visits with multiple types of disease were also removed, so to rule out multi-label cases. The ICD-10 codes of these diseases are specified in Table 4. Lastly, visit pairs (*v*_1_, *v*_2_) with different labels were remained. We used the average of code embeddings in *v*_1_ to predict the disease label of *v*_2_.

#### 4.1.2 Mortality Prediction

MIMIC-IV includes 21,622 ICU patients having diagnostic, procedures, and medication records. We collected the set of visits of these patients within a month since they entered the ICU, and unwrapped the set of visits into a set of medical codes. Each input is a set of medical codes of an ICU patient, and the task is to predict the label, which is 0/1 (denoting deceased or live within a month). As the positive samples, which are labelled as 1, are far more than the negative samples, we randomly down-sampled positive samples to the same size of the negative samples.

#### 4.1.3 Medication Recommendation

This task used the diagnostic/procedure information of a visit to predict the probability that each NDC would be in the prescription for this visit. To get the features of a visit, we firstly embedded the diagnostic/procedures codes in this visit and each prescription code for this visit using the pretrained embedding from the same kind of model. Then we concatenated the averaged embedding of diagnostic/procedures embeddings and the embedding for one of the prescription codes as the feature of a sample with the training label as 1.

### 4.2 Results

The evaluation results are shown in the Table. 2 and Table. 3.

**Table 2.** Evaluation results on diagnoses data. Model name with ‘(v)’ means pre-training with the visit-level data. For classification tasks, we used accuracy, macro-precision, macro-recall, and macro-F1 as the metrics. To evaluate the regression task, we used explained variance and mean-squared error (MSE), where higher explained variance and lower MSE is better. We ran each experiment 5 times, and all of the standard deviations were less than 2*e* − 2.

|  | Next Visit<br>Diagnoses Prediction |  |  |  | Mortality<br>Prediction |  |  |  | Medication<br>Recommendation |  |
| --- | --- | --- | --- | --- | --- | --- | --- | --- | --- | --- |
|  | Accuracy | Precision | Recall | F1 | Accuracy | Precision | Recall | F1 | Explained Variance | MSE |
| BERT | 0.547 | 0.470 | 0.386 | 0.406 | 0.659 | 0.653 | 0.653 | 0.652 | 0.783 | 0.054 |
| GloVe | 0.632 | 0.598 | 0.520 | 0.548 | 0.693 | 0.695 | 0.695 | 0.693 | **0.806** | **0.048** |
| GloVe (v) | **0.632** | **0.598** | **0.522** | **0.551** | **0.694** | **0.696** | **0.696** | **0.694** | 0.805 | 0.049 |
| Word2Vec | 0.515 | 0.427 | 0.354 | 0.368 | 0.648 | 0.655 | 0.651 | 0.650 | 0.760 | 0.060 |
| Word2Vec (v) | 0.514 | 0.491 | 0.332 | 0.342 | 0.650 | 0.654 | 0.653 | 0.650 | 0.759 | 0.060 |
| LDA | 0.558 | 0.514 | 0.369 | 0.389 | 0.664 | 0.668 | 0.667 | 0.663 | 0.581 | 0.105 |
| LDA (v) | 0.548 | 0.506 | 0.362 | 0.388 | 0.658 | 0.664 | 0.662 | 0.658 | 0.607 | 0.098 |
| random | 0.510 | 0.395 | 0.307 | 0.311 | 0.657 | 0.660 | 0.660 | 0.657 | 0.759 | 0.060 |

**Table 3.** Evaluation results on procedures and medications data.

|  |  | Procedures |  |  |  |  |  |  |  | Medications |  |  |  |  |  |  |  |
| --- | --- | --- | --- | --- | --- | --- | --- | --- | --- | --- | --- | --- | --- | --- | --- | --- | --- |
|  |  | BERT | GloVe | GloVe (v) | Word-2Vec | Word2-Vec (v) | LDA | LDA (v) | random | BERT | GloVe | GloVe (v) | Word-2Vec | Word2-Vec (v) | LDA | LDA (v) | random |
| Mortality Prediction | Accuracy | 0.668 | **0.68** | 0.678 | 0.647 | 0.648 | 0.659 | 0.639 | 0.659 | 0.621 | 0.648 | **0.65** | 0.623 | 0.62 | 0.632 | 0.636 | 0.62 |
|  | Precision | 0.669 | **0.681** | 0.661 | 0.647 | 0.648 | 0.66 | 0.642 | 0.659 | 0.623 | 0.649 | **0.652** | 0.628 | 0.622 | 0.634 | 0.638 | 0.62 |
|  | Recall | 0.668 | **0.68** | 0.678 | 0.647 | 0.648 | 0.659 | 0.64 | 0.659 | 0.623 | 0.649 | **0.652** | 0.622 | 0.622 | 0.634 | 0.638 | 0.618 |
|  | F1 | 0.668 | **0.68** | 0.678 | 0.647 | 0.648 | 0.658 | 0.639 | 0.659 | 0.62 | 0.647 | **0.65** | 0.618 | 0.62 | 0.631 | 0.636 | 0.617 |
| Medication Recommendation | Explained Variance | 0.780 | **0.813** | 0.803 | 0.787 | 0.788 | 0.757 | 0.76 | 0.787 | / |  |  |  |  |  |  |  |
|  | MSE | 0.055 | **0.047** | 0.049 | 0.053 | 0.053 | 0.061 | 0.06 | 0.053 |  |  |  |  |  |  |  |  |

Table. 2 shows that all four methods have improved performance in diagnosis prediction compared with the baseline, randomly initialized embeddings. It indicates that the hidden information captured by these models has predictive powers for diagnoses. While in Table. 3, for procedures, the embeddings learned by LDA and Word2Vec did worse than the baseline. From Table. 2 and Table. 3, for the ICU patients mortality prediction, we can see that the diagnosis embeddings pretrained by GloVe are more explicit in showing the patient’s criticality in syndromes compared to procedures and medications data. While in the medication recommendation task, only embeddings learned by GloVe performed better than the randomly initialized inputs both for diagnoses and procedures. The distribution biases introduced by LDA could worsen the medication recommendation results. It may be caused by the poor interpretability of the LDA pre-trained embeddings of NDC codes.

Comparing the granularity of pre-training data, using pre-training data at the patient-level is better than using the visit-level for co-occurrence-based methods (GloVe, Word2Vec, and LDA) in the diagnoses prediction task. Pre-training with the visit-level data may lose the temporal relationship between diseases, especially for chronic diseases. However, for mortality prediction, as the critical factor is mainly determined by the fetal diagnoses [18] such as respiratory failure and sepsis, which is acute and often unexpected, there is no big difference between using the two kinds of data granularities. It is worth noting that GloVe showed the best performance for all three downstream tasks among all models considered. One of the reasons that GloVe outperformed BERT may be that BERT was not fine-tuned for these tasks, which we deliberately left out since we wanted to make the embeddings as generalized as possible. Embeddings learned by Word2Vec and LDA performed poorly on these tasks. Word2Vec does not use the global co-occurrence information, instead, it uses the context information within a fixed-size window; LDA is designed to do topic modelling. Nevertheless, embeddings learned by LDA have better performance than the ones learned by Word2Vec in the diagnoses prediction task and mortality prediction task.

## 5 DISCUSSION

Surprisingly, BERT did not do well in the evaluation. The reason may be that BERT is only pre-trained with MLM. Besides, the pre-training of BERT is sensitive to hyper-parameters and needs a large corpus, so the optimized pre-training method RoBERTa [24] can further be utilized for medical concepts representation learning.

With Word2Vec, the fixed window size introduces biases induced by the order of the NDC codes, which might be the main reason for the poor performances of the Word2Vec embeddings. In other words, the neighborhood of a medical code is restrained by the window size, leading to the omitting of codes that are out of the window but in the same visit.

For the tasks like clinical concept extraction in biomedical text, some published work has shown that BERT is better than GloVe and Word2Vec [39, 1, 32]. But in EHR data, Getzen et al. [14] also finds that BERT did not perform well when using static embeddings. Combining the results of this paper, we find that longitudinal EHR data is context-independent within a visit (i.e. the position of disease in the visit does not impact the semantics) but is context-dependent between visits. GloVe and Word2Vec word embeddings are context-independent, while BERT generates different embeddings for a word to capture the context of the word, which is context-dependent. So BERT can introduce disturbance for medical code embeddings as there is no context but co-occurrence within a visit. And it is more suitable to use context-independent models e.g. GloVe and Word2Vec when using the visit as the sample for word embedding.

## 6 CONCLUSION

We have presented a new perspective on medical concepts representation learning methods comparison via visualizations and three downstream tasks. We considered three kinds of medical concepts: ICD-10-CM (diagnoses), ICD-10-PCS (procedures), and NDC (medications) and four pre-training methods — LDA, Word2Vec, GloVe, and BERT using the MIMIC-IV dataset. We compare the effectiveness of four methods in embedding three modalities of clinical codes. BERT pre-trained embeddings without fine-tuning cannot beat the embeddings learned by GloVe in this comparison framework. Furthermore, interestingly, embeddings learned by LDA are better than Word2Vec in the next visit diagnoses prediction task and mortality prediction task. Considering the visualizations and the results of the downstream tasks, pre-training with GloVe is the best in capturing the hidden semantics between medical codes in MIMIC-IV. Moreover, the granularity of pre-training samples should be considered according to different applications. In the mortality prediction, using visit-level data could be better, while in the medication recommendation, using patient-level samples may be a better choice.

## Data Availability

All data produced in the present study are available upon reasonable request to the authors

https://bit.ly/3ONj9Su

## 7 ACKNOWLEDGMENT

## 8 SUPPLEMENTARY MATERIAL

The classified ICD-10-CM codes in Figure. 3 are specified according to Table. 4.

**Table 4.** Specified ICD-10 code prefix for six types diseases. We use the prefix to denote all of the codes with this prefix. The support is the occurrence amount of the codes in the specified class. The classification of codes refers to [20, 12].

| Diseases | ICD-10 codes | Num. of codes | Support |
| --- | --- | --- | --- |
| Chronic kidney failure | N18 | 7 | 26507 |
| Acute kidney failure | N17,O904,T8612,N990 | 8 | 24737 |
| Respiratory failure | J960,J962,J969,J9582,P285 | 12 | 11917 |
| Heart failure | I50, I9713,I0981, I110,I130,I132 | 29 | 44832 |
| Stroke | H341,I63,I64,I61,G45 | 101 | 5637 |
| Diabetes | E10,E11,E12,E13,E14 | 158 | 63027 |

## 9 DATA AVAILABILITY

The MIMIC-IV data underlying this article is available in https://doi.org/10.13026/a3wn-hq05 with restricted-access.

## Footnotes

1 https://bit.ly/3ONj9Su

2 https://www.icd10data.com/ICD10CM/Codes

3 https://www.cms.gov/medicare/icd-10/2021-icd-10-pcs

